# Five-Year Breast Cancer Risk Prediction From Screening Breast Ultrasound Using Deep Learning

**DOI:** 10.64898/2026.06.21.26356188

**Authors:** Yuxuan Chen, Haoyan Yang, Yanqi Xu, Ria Soni, Laura Heacock, Maciej Lis, Aleksandra Stanek, Tomasz Puto, Alana Amarosa Lewin, Linda Moy, Freya Schnabel, Yiqiu Shen

## Abstract

**Objective:** To develop and evaluate a deep learning model for five-year breast cancer risk prediction from screening breast ultrasound (BUS) examinations.

**Methods:** This retrospective study included 295,298 breast ultrasound examinations from 122,072 women imaged between 2012 and 2020. Patients were split into training, validation, and test sets; the test set included screening examinations only. BUS-Risk-Net aggregated image features using attention-based multiple instance learning and combined them with age and ultrasound-estimated breast density to predict 2- to 5-year risk. Performance was compared with the full Tyrer–Cuzick model in a matched case-control cohort and with a reduced Tyrer–Cuzick model in the held-out test set. Risk stratification was evaluated within BI-RADS density categories.

**Results:** In the matched case-control cohort (n = 240 women), BUS-Risk-Net achieved a 5-year AUC of 0.632 (95% CI, 0.562–0.702), versus 0.514 for the full Tyrer–Cuzick model (95% CI, 0.440–0.588; *p* = 0.04). Among 19,548 examinations from 9,015 women eligible for 5-year evaluation in the test set, BUS-Risk-Net achieved an AUC of 0.679 (95% CI, 0.653–0.706), versus 0.594 for the reduced Tyrer–Cuzick model (95% CI, 0.564–0.623; *P* < .001). Observed 5-year cancer incidence increased across AI-defined risk tiers within each BI-RADS density category, ranging from 0.0% to 5.8% after AI stratification, compared with 2.1% to 3.6% across density categories alone.

**Discussion:** Deep learning models applied to screening breast ultrasound could enable long-term breast cancer risk prediction and stratify risk beyond breast density alone. External and prospective validation is needed before clinical use.

**Summary Sentence:** BUS-Risk-Net enabled long-term breast cancer risk prediction from screening ultrasound, outperformed the Tyrer–Cuzick–based comparators, and identified distinct 5-year risk groups with differing observed cancer incidence within each breast density category.

## Introduction

Breast cancer is the leading cause of cancer-related deaths among women worldwide, with an estimated 2.3 million new cases and 670,000 deaths in 2022^1^. Unlike breast cancer screening, which aims to detect existing malignancy, risk prediction estimates the likelihood that a cancer-free individual will develop breast cancer over a defined future period. Accurate risk prediction is clinically important because it can guide decisions about supplemental screening, chemoprevention, and risk-reducing surgery^2–4^.

Current risk prediction relies primarily on clinical models such as the Breast Cancer Risk Assessment Test (Gail), Tyrer–Cuzick, and the Breast Cancer Surveillance Consortium model^5–8^. While widely adopted, these models achieve only moderate discriminatory performance, with reported 5-year AUCs of approximately 0.6^9^ despite the inclusion of mammographic breast density. Deep learning models trained on mammograms have improved breast cancer risk prediction compared with conventional clinical risk models by capturing risk-relevant imaging features, such as calcifications, localized bilateral dissimilarity, and other subtle patterns not fully captured by clinical variables^10–14^.

However, mammography-derived risk assessment has specific limitations and may be impractical in settings where mammography is unavailable. In low-resource settings, screening mammography is difficult to obtain because it requires expensive equipment, trained personnel, and quality-assurance infrastructure^15,16^. In populations where breast cancer diagnosis peaks at younger ages and breast density is higher, including many Asian populations, mammography may also have reduced sensitivity^17–19^. Breast ultrasound, which is accessible, relatively low cost, radiation-free, and well suited for younger women, pregnant women, and women with dense breasts, is widely used as a complementary or alternative screening modality in these settings^20,21^. These features make ultrasound a practical imaging source for risk prediction when mammography is unavailable, limited, or performed at staggered intervals.

Prior studies suggest that breast ultrasound contains imaging biomarkers associated with future breast cancer risk independent of mammographic density, including glandular tissue composition and background echotexture patterns^22,23^. Despite this evidence, ultrasound-based risk prediction models remain largely unexplored. Existing work has focused on estimating breast density from ultrasound for incorporation into existing clinical risk models^24^. This approach may not capture the broader range of risk-relevant information contained in ultrasound images.

To address this gap, we developed the breast ultrasound–based risk prediction network (BUS-Risk-Net), a deep learning framework that predicts breast cancer risk from breast ultrasound screening exams. We hypothesized that breast ultrasound images encode risk-relevant information that can be learned by a deep learning model. The aim of this study was to develop and evaluate an ultrasound-based deep learning model that provides individualized breast cancer risk estimates in screening settings where mammography is unavailable, limited, or not routinely used.

## Materials and Methods

This retrospective study was approved by the Institutional Review Board, with waiver of informed consent. This study followed the Checklist for Artificial Intelligence in Medical Imaging (CLAIM) guideline^25^.

### Data Collection

The dataset construction process is illustrated in Figure 1. We retrospectively identified a consecutive cohort of female adult (>18 years old) patients who underwent breast ultrasound at Institution A between January 1, 2012, and December 31, 2020, including both screening and diagnostic imaging. The initial cohort comprised 414,016 examinations from 185,054 patients.

**Figure 1.**
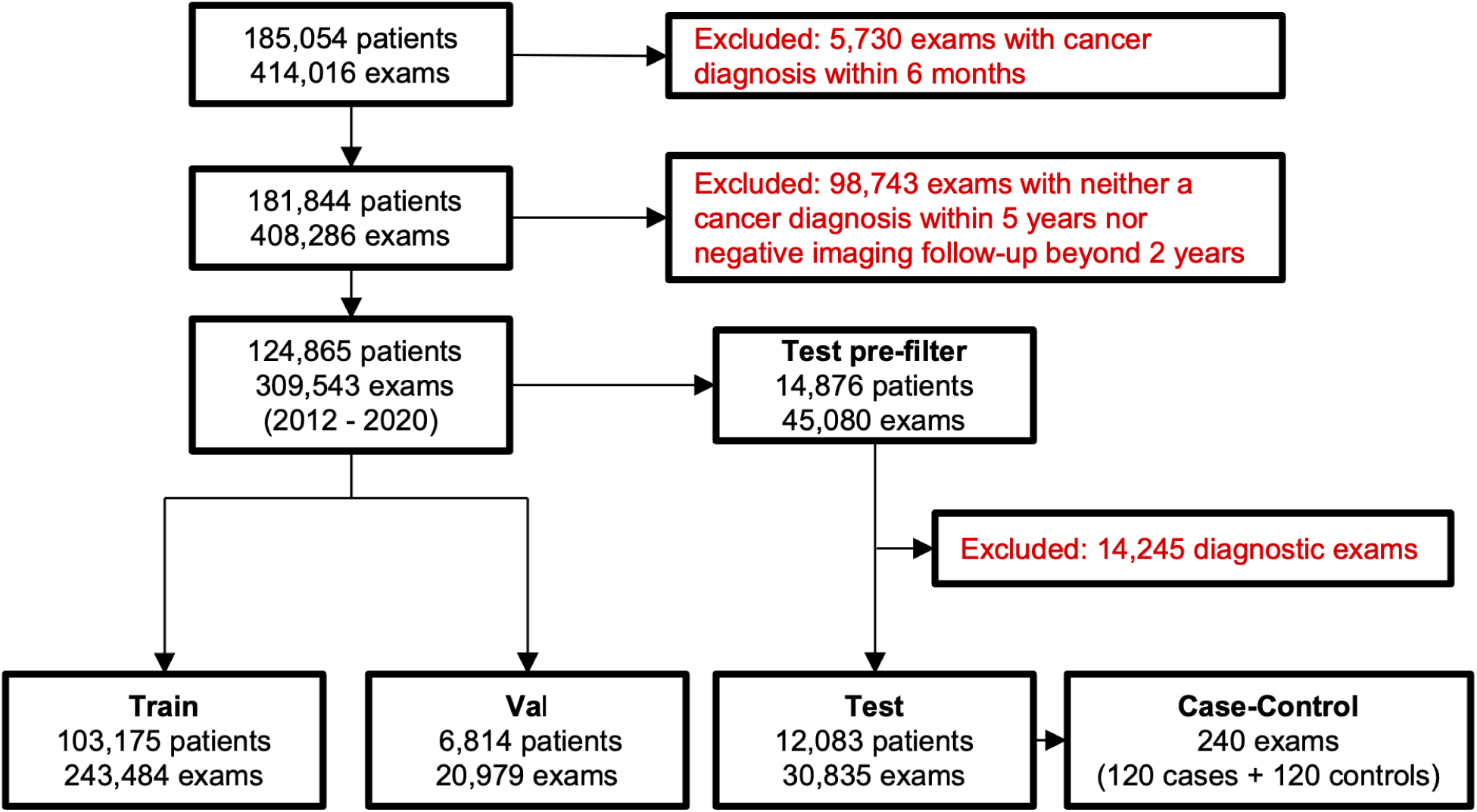
Flowchart of cohort selection and dataset partitioning.

Two exclusion steps were applied sequentially. First, we excluded examinations associated with a biopsy-confirmed malignancy within 6 months of imaging (n = 5,730), because these cancers were likely present at the time of ultrasound and were not considered future risk events. Second, we excluded examinations that had neither biopsy-confirmed breast cancer within 5 years nor at least 2 years of negative imaging follow-up (n = 98,743). This ensured that each included examination had sufficient follow-up to contribute to at least one prediction horizon from 2 to 5 years (see Outcome Definition).

Patients were randomly split at the patient level into training, validation, and test sets, with no patient appearing in more than one partition. The training set included 243,484 examinations from 103,175 patients, and the validation set included 20,979 examinations from 6,814 patients. Training and validation included both screening and diagnostic examinations to increase imaging diversity. Because the intended use was future risk prediction at routine screening, the held-out test set was restricted to screening examinations only. This strategy has been adopted in the literature^26–28^. From 45,080 test-set examinations in 14,876 patients, we excluded 14,245 diagnostic examinations, resulting in a final screening test set of 30,835 examinations from 12,083 patients. The number of evaluable examinations differed by prediction horizon because negative examinations required follow-up beyond the corresponding horizon.

A matched case-control cohort was sampled from the screening test set to compare BUS-Risk-Net with the full Tyrer–Cuzick model^8^. This cohort included 240 patients: 120 cases and 120 controls. Cases were screening examinations from women who were cancer-free at imaging and developed biopsy-confirmed breast cancer within 5 years. Controls were screening examinations from women who remained cancer-free for at least 5 years. Matching was performed by age group in 10-year strata, race, and breast density, categorized as non-dense breasts according to the Breast Imaging Reporting and Data System (BI-RADS A–B) or dense breasts (BI-RADS C–D).

### Outcome Definition

Pathology records from 2010 to 2025 were used as the reference standard for breast cancer outcome and timing. Malignant outcomes included invasive ductal carcinoma, invasive lobular carcinoma, ductal carcinoma in situ, microinvasive carcinoma, inflammatory carcinoma, intraductal papillary carcinoma, and special histologic subtypes, including tubular, mucinous, and cribriform carcinoma. Separate labels were assigned for 2-, 3-, 4-, and 5-year prediction horizons. For a given horizon, an examination was labeled positive if pathology confirmed breast cancer within that interval after the examination date. Event time was defined as the interval from the examination date to the first positive pathology result. An examination was labeled negative for a given horizon if no malignancy was identified and follow-up extended beyond that horizon. Negative follow-up was defined as either subsequent negative imaging assessments (BI-RADS 1 or 2) or abnormal findings (BI-RADS 0, 3, 4, or 5) that were subsequently proven benign on pathology or the follow-up exam. Examinations without malignant outcomes were censored at the last documented negative follow-up.

### Image Preprocessing

Image preprocessing followed prior work^29,30^. Raw ultrasound frames were extracted from the PixelData field of DICOM files. Images were cropped to retain the breast field of view and remove peripheral annotations and background. To reduce variation across ultrasound systems and scan geometries, the pipeline localized the acoustic region and generated standardized rectangular crops. The mean crop size was 609.4 × 674.7 pixels.

### Model Development

BUS-Risk-Net was designed to predict each of 2- to 5-year future breast cancer risk from breast ultrasound examinations by combining imaging features with age and ultrasound-estimated breast density based on the amount of fibroglandular breast tissue^31^ (Figure 2). The model included an imaging stream and a clinical stream. The imaging stream extracts visual features from ultrasound images, while the clinical stream encodes patient-level risk factors; the two representations are fused to produce a final risk estimate.

**Figure 2.**
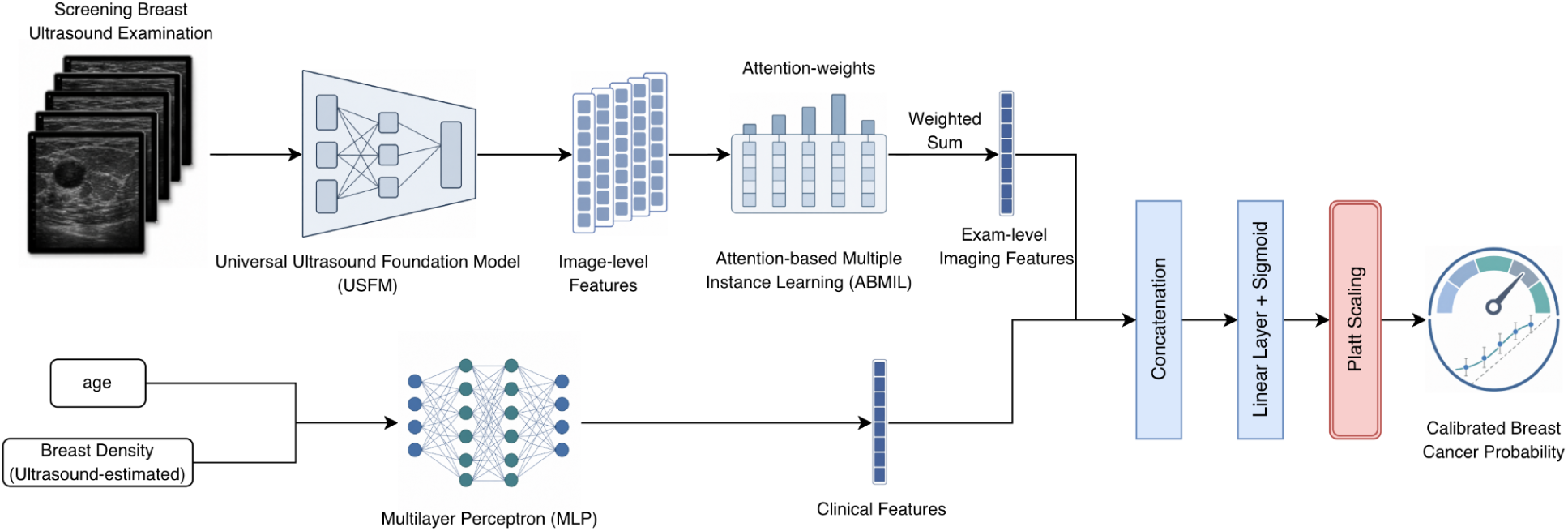
Schematic diagram of breast ultrasound–based risk prediction network (BUS-Risk-Net). Breast ultrasound images from a single exam are processed by the universal ultrasound foundation model (USFM) and aggregated via an attention-based multiple instance learning (ABMIL) module. Clinical risk factors (age and ultrasound-estimated breast density) are encoded through a multilayer perceptron (MLP). The image and clinical features are concatenated, passed through a linear layer with sigmoid activation, and calibrated via Platt scaling to produce the calibrated breast cancer risk probability.

In the imaging stream, each ultrasound image was encoded using the universal ultrasound foundation model (USFM)^32^, which was pretrained on more than 3 million ultrasound images. Because the number of images varied across examinations, attention-based multiple instance learning (ABMIL) was used to aggregate image-level features into one examination-level imaging representation^33^. In the clinical stream, age and ultrasound-estimated breast density were encoded using a multilayer perceptron. Breast density was estimated directly from ultrasound images using a previously described method^24^. The imaging and clinical representations were concatenated and passed through a final prediction layer to generate a risk score.

Separate models were trained for 2-, 3-, 4-, and 5-year prediction horizons. Models were optimized using AdamW with weighted cross-entropy loss for 20 epochs. Learning rates were sampled from a log-uniform distribution between 4 × 10^-5 and 6 × 10^-5. Model selection was based on validation-set AUC. For final testing, predictions were averaged across the three models with the highest validation AUCs. Predicted probabilities were calibrated using Platt scaling^34^ fit on the validation set.

### Model Performance Comparisons

BUS-Risk-Net was compared with the Tyrer–Cuzick model, version 8, using two complementary approaches. This strategy was used because several Tyrer–Cuzick input variables, including family history, reproductive history, prior biopsy results, and hormone use, were not routinely available in structured electronic health record fields. The Tyrer–Cuzick model was accessed through the IBIS Breast Cancer Risk Evaluation Tool^35^.

First, we compared BUS-Risk-Net with the full Tyrer–Cuzick model in the matched case-control cohort. Tyrer–Cuzick risk factors were obtained by structured chart review performed by a resident physician (R.S.) and supervised by an attending breast radiologist (L.H.) with 9 years of experience. Investigators were blinded to outcome status during chart review.

Second, we compared BUS-Risk-Net with a reduced Tyrer–Cuzick variant on the full held-out screening test set^24^. This reduced Tyrer–Cuzick model used age and ultrasound-estimated breast density as inputs, while all other Tyrer–Cuzick variables were assigned the model’s predefined missing-value placeholders.

We also performed an ablation analysis using an image-only version of BUS-Risk-Net. This model excluded age and breast density and was used to assess the independent predictive value of ultrasound imaging features.

### Risk Stratification Analysis

Following prior studies^36,37^, the predicted 5-year risk was stratified into three risk tiers: average risk (<1.7%), intermediate risk (1.7%–3.0%), and high risk (≥3.0%). These thresholds were derived from NCCN^38^ and ASCO^39^ guidelines. They were used to assess risk stratification within the study cohort. To determine whether BUS-Risk-Net provided information beyond breast density, screening test-set examinations were grouped by mammography-based BI-RADS density category and AI-predicted risk tier. Observed 5-year breast cancer incidence was then calculated within each subgroup.

### Statistical Analysis

Model discrimination was evaluated using AUC for 2-, 3-, 4-, and 5-year prediction horizons. Ninety-five percent confidence intervals were estimated using 5,000 bootstrap resamples performed at the patient level to account for correlation among multiple examinations from the same patient. Pairwise AUC comparisons were performed using the DeLong test for correlated receiver operating characteristic curves. Statistical significance was defined as a two-sided *p* value less than 0.05. Calibration was evaluated in the held-out screening test set using the Brier score, expected calibration error, calibration slope, and calibration intercept. Calibration plots compared predicted probabilities with observed event rates across deciles of predicted risk. Subgroup analyses were performed at the exam level across age groups, BI-RADS breast density categories, and racial groups for the 5-year prediction horizon.

## Results

The final study cohort included 122,072 women who underwent 295,298 breast ultrasound examinations at Institution A between 2012 and 2020. Among these examinations, 5,580 (1.9%) were followed by a breast cancer diagnosis within 5 years. The mean time to diagnosis was 3.3 ± 1.1 years. Demographic and clinical characteristics of the overall cohort are summarized in Table 1. Characteristics of the matched case-control cohort are provided in Supplementary Table S1. The distribution of time to diagnosis and the number of examinations evaluated at each prediction horizon are provided in Supplementary Tables S2 and S3.

**Table 1.** Cohort characteristics across the full dataset and training, validation, and test splits. “5-year positive exams” denotes examinations followed by a breast cancer diagnosis within 5 years; “5-year positive patients” denotes patients with at least one 5-year positive exam. SD = standard deviation; BI-RADS = Breast Imaging Reporting and Data System.

| Characteristics | All | Train | Val | Test |
| --- | --- | --- | --- | --- |
| **Patient-level** |  |  |  |  |
| All patients | 122,072 (100%) | 103,175 (100%) | 6,814 (100%) | 12,083 (100%) |
| 5-year positive patients | 2,993 (2.5%) | 2,387 (2.3%) | 217 (3.2%) | 389 (3.2%) |
| **Race** |  |  |  |  |
| White | 76,108 (62.3%) | 63,388 (61.4%) | 4,543 (66.7%) | 8,177 (67.7%) |
| African American | 11,758 (9.6%) | 10,182 (9.9%) | 587 (8.6%) | 989 (8.2%) |
| Asian | 5,408 (4.4%) | 4,724 (4.6%) | 245 (3.6%) | 439 (3.6%) |
| Hispanic | 1,713 (1.4%) | 1,458 (1.4%) | 98 (1.4%) | 157 (1.3%) |
| Unknown | 27,085 (22.2%) | 23,423 (22.7%) | 1,341 (19.7%) | 2,321 (19.2%) |
| **Exam-level** |  |  |  |  |
| All examinations | 295,298 (100%) | 243,484 (100%) | 20,979 (100%) | 30,835 (100%) |
| 5-year positive exams | 5,580 (1.9%) | 4,500 (1.8%) | 438 (2.1%) | 642 (2.1%) |
| Time to diagnosis, years, mean (SD) | 3.3 (1.1) | 3.3 (1.1) | 3.3 (1.1) | 3.4 (1.1) |
| **Age** |  |  |  |  |
| <40 | 18,188 (6.2%) | 17,061 (7.0%) | 615 (2.9%) | 512 (1.7%) |
| 40–49 | 76,831 (26.0%) | 64,928 (26.7%) | 5,161 (24.6%) | 6,742 (21.9%) |
| 50–59 | 91,322 (30.9%) | 74,648 (30.7%) | 6,665 (31.8%) | 10,009 (32.5%) |
| 60–69 | 70,532 (23.9%) | 56,168 (23.1%) | 5,543 (26.4%) | 8,821 (28.6%) |
| ≥70 | 38,425 (13.0%) | 30,679 (12.6%) | 2,995 (14.3%) | 4,751 (15.4%) |
| **Density** |  |  |  |  |
| A – Fatty | 5,346 (1.8%) | 4,583 (1.9%) | 271 (1.3%) | 492 (1.6%) |
| B – Scattered | 74,959 (25.4%) | 61,738 (25.4%) | 5,260 (25.1%) | 7,961 (25.8%) |
| C – Heterogeneous | 177,506 (60.1%) | 145,165 (59.6%) | 13,034 (62.1%) | 19,307 (62.6%) |
| D – Dense | 31,236 (10.6%) | 26,006 (10.7%) | 2,225 (10.6%) | 3,005 (9.7%) |
| Unknown | 6,251 (2.1%) | 5,992 (2.5%) | 189 (0.9%) | 70 (0.2%) |
| **BI-RADS** |  |  |  |  |
| 0 — incomplete assessment | 15,527 (5.3%) | 12,540 (5.2%) | 977 (4.7%) | 2,010 (6.5%) |
| 1/2/3 — negative or benign | 266,798 (90.3%) | 219,058 (90.0%) | 19,134 (91.2%) | 28,606 (92.8%) |
| 4 — suspicious abnormality | 12,973 (4.4%) | 11,886 (4.9%) | 868 (4.1%) | 219 (0.7%) |
| **Device** |  |  |  |  |
| Siemens | 132,419 (44.8%) | 111,084 (45.6%) | 9,020 (43.0%) | 12,315 (39.9%) |
| Philips | 132,427 (44.8%) | 106,922 (43.9%) | 9,708 (46.3%) | 15,797 (51.2%) |
| GE | 14,462 (4.9%) | 11,944 (4.9%) | 978 (4.7%) | 1,540 (5.0%) |
| Toshiba | 10,272 (3.5%) | 8,985 (3.7%) | 798 (3.8%) | 489 (1.6%) |
| Medison | 5,703 (1.9%) | 4,535 (1.9%) | 475 (2.3%) | 693 (2.2%) |
| Other | 15 (0.0%) | 14 (0.0%) | 0 (0.00%) | 1 (0.00%) |

### Comparison Against the Full Tyrer–Cuzick Model in the Case–Control Cohort

In the matched case-control cohort, BUS-Risk-Net had a higher 5-year AUC than the full Tyrer–Cuzick model with complete clinical risk factors (0.632; 95% CI, 0.562–0.702 vs 0.514; 95% CI, 0.440–0.588; *p* = 0.04) (Table 2). ROC curves are shown in Figure 3.

**Figure 3.**
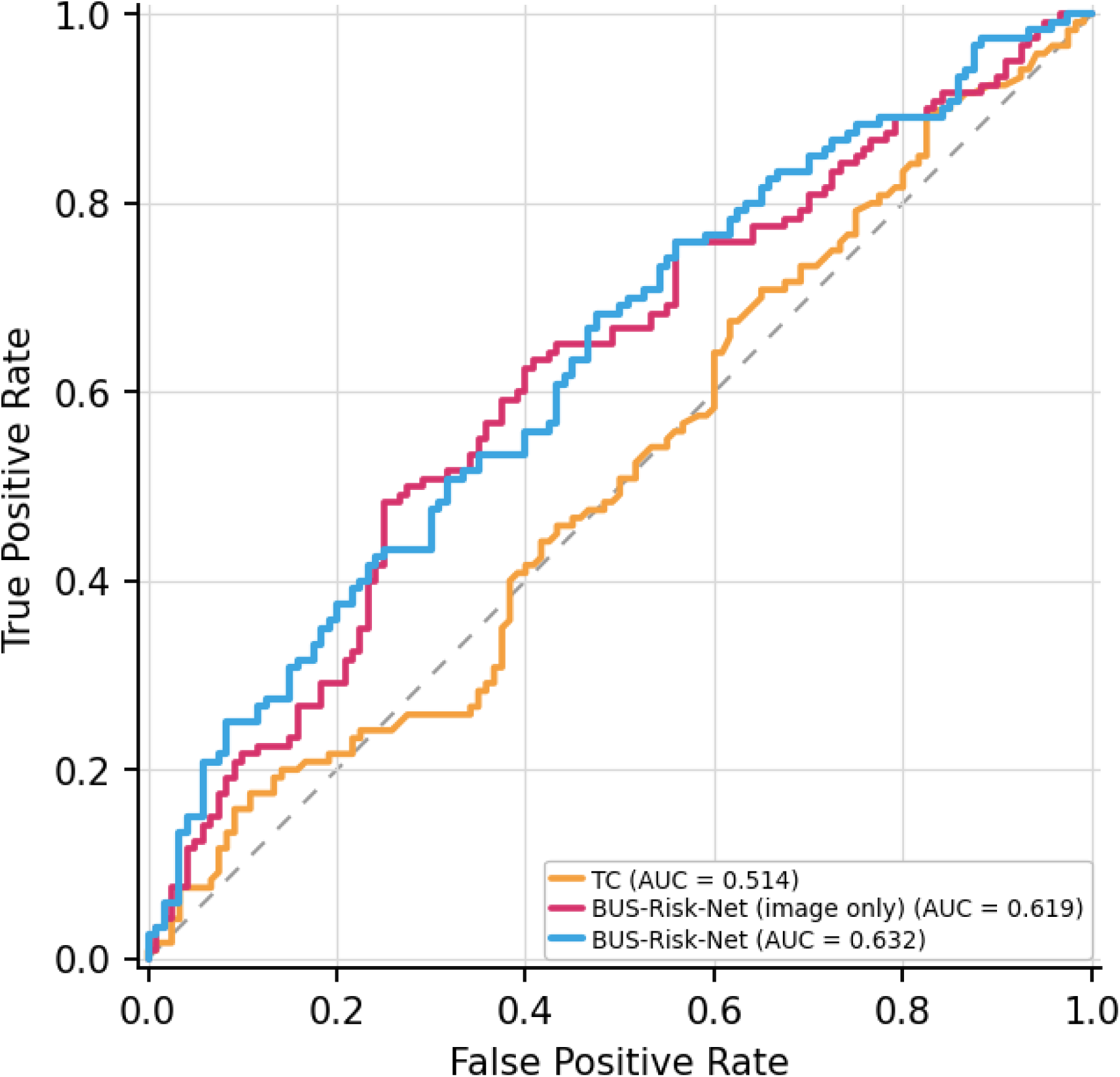
Receiver operating characteristic (ROC) curves for 5-year breast cancer risk prediction in the matched case–control cohort (n = 240) for breast ultrasound–based risk prediction network (BUS-Risk-Net), the image-only variant of BUS-Risk-Net, and the full Tyrer–Cuzick (TC) model with all risk factors.

**Table 2.** Performance of breast ultrasound–based risk prediction network (BUS-Risk-Net) and comparator models for 2-, 3-, 4-, and 5-year breast cancer risk prediction. Areas under the receiver operating characteristic curve (AUCs) with 95% confidence intervals (CIs) are reported in the held-out test set for the reduced Tyrer–Cuzick (TC) variant, image-only BUS-Risk-Net, and full BUS-Risk-Net. For 5-year prediction, performance of the full TC model was additionally evaluated in the case–control cohort. *P* values were calculated using the DeLong test, comparing each alternative model with the full BUS-Risk-Net model, which served as the reference (ref). Dashes indicate that the model was not evaluated in the corresponding cohort or prediction interval.

| Method | 2-Year (Held-out Test) |  | 3-Year (Held-out Test) |  | 4-Year (Held-out Test) |  | 5-Year (Held-out Test) |  | 5-Year (Case–Control) |  |
| --- | --- | --- | --- | --- | --- | --- | --- | --- | --- | --- |
|  | AUC | *p* | AUC | *p* | AUC | *p* | AUC | *p* | AUC | *p* |
| Full TC model | — | — | — | — | — | — | — | — | 0.514<br>(0.440–0.588) | 0.04 |
| Reduced TC Variant | 0.636<br>(0.564–0.706) | 0.006 | 0.615<br>(0.574–0.655) | 0.002 | 0.617<br>(0.582–0.650) | < 0.001 | 0.594<br>(0.564–0.623) | < 0.001 | — | — |
| BUS-Risk-Net (image only) | 0.692<br>(0.635–0.746) | 0.003 | 0.639<br>(0.605–0.674) | 0.02 | 0.631<br>(0.598–0.662) | < 0.001 | 0.648<br>(0.621–0.676) | < 0.001 | 0.619<br>(0.548–0.690) | 0.61 |
| **BUS-Risk-Net** | **0.741**<br>**(0.689–0.790)** | ref | **0.669**<br>**(0.634–0.702)** | ref | **0.665**<br>**(0.634–0.695)** | ref | **0.679**<br>**(0.653–0.706)** | ref | **0.632**<br>**(0.562–0.702)** | ref |

### Performance on the Held-Out Screening Test Set

On the held-out screening test set (Table 2), BUS-Risk-Net had higher AUCs than the reduced Tyrer–Cuzick variant at all prediction horizons^24^. BUS-Risk-Net achieved AUCs of 0.741 (95% CI, 0.689–0.790) at 2 years, 0.669 (95% CI, 0.634–0.702) at 3 years, 0.665 (95% CI, 0.634–0.695) at 4 years, and 0.679 (95% CI, 0.653–0.706) at 5 years. The corresponding AUCs for the reduced Tyrer–Cuzick variant were 0.636 (95% CI, 0.564–0.706; *p* = 0.006), 0.615 (95% CI, 0.574–0.655; *p* = 0.002), 0.617 (95% CI, 0.582–0.650; *p* < 0.001), and 0.594 (95% CI, 0.564–0.623; *p* < 0.001). The largest difference was observed at the 2-year horizon, with a ΔAUC of 0.105. ROC curves are shown in Figure 4.

**Figure 4.**
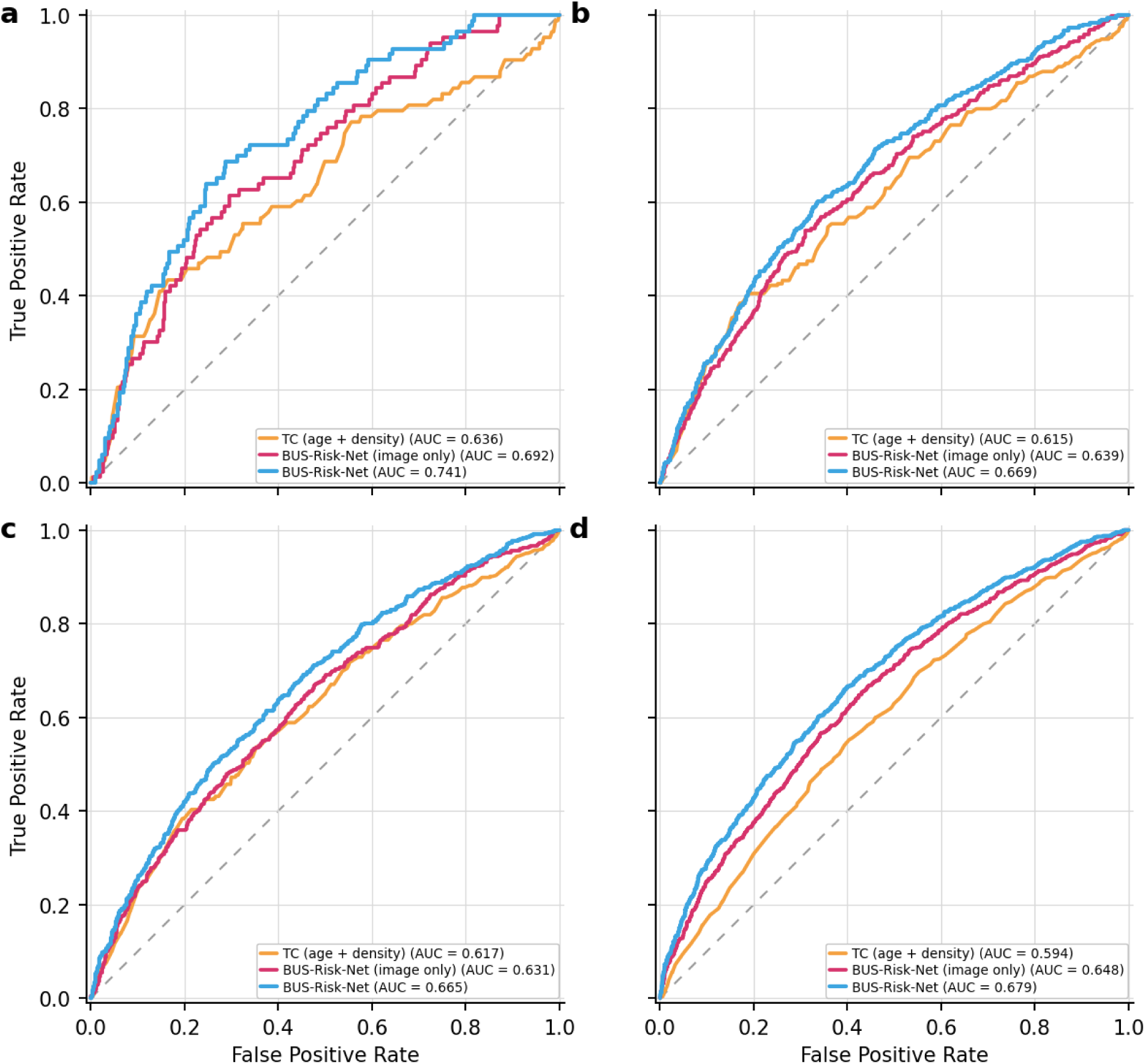
Receiver operating characteristic (ROC) curves for 2- to 5-year breast cancer risk prediction in the held-out test set for breast ultrasound–based risk prediction network (BUS-Risk-Net), the image-only variant of BUS-Risk-Net, and the reduced Tyrer–Cuzick (TC) variant incorporating age and ultrasound-estimated breast density.

### Ablation Study on Image-only Variant

The image-only variant of BUS-Risk-Net excluded age and ultrasound-estimated breast density. In the matched case-control cohort, the image-only model achieved a 5-year AUC of 0.619 (95% CI, 0.548–0.690), compared with 0.514 (95% CI, 0.440–0.588) for the full Tyrer–Cuzick model (Table 2). On the held-out screening test set, the image-only model achieved AUCs of 0.692 (95% CI, 0.635–0.746) at 2 years, 0.639 (95% CI, 0.605–0.674) at 3 years, 0.631 (95% CI, 0.598–0.662) at 4 years, and 0.648 (95% CI, 0.621–0.676) at 5 years. These AUCs were higher than those of the reduced Tyrer–Cuzick variant at each horizon but lower than those of the full BUS-Risk-Net model (Table 2).

### Calibration

Calibration of 5-year risk predictions on the held-out screening test set is shown in Supplementary Figure 1. Predicted probabilities were aligned with observed event rates across deciles of predicted risk. The Brier score was 0.031 (95% CI, 0.028–0.034). Expected calibration error was 0.002 (95% CI, 0.001–0.006). Calibration slope was 1.053 (95% CI, 0.884–1.235). Calibration intercept was 0.067 (95% CI, −0.052–0.176).

### Density-Stratified Risk Stratification

BUS-Risk-Net provided 5-year risk stratification within each BI-RADS breast density category (Table 3). Across density groups, observed cancer incidence was higher in the AI-defined high-risk group than in the average-risk group: 5.3% versus 0.0% for fatty breasts, 4.3% versus 1.3% for scattered fibroglandular breasts, 5.8% versus 1.5% for heterogeneously dense breasts, and 4.3% versus 0.9% for extremely dense breasts. The distribution of AI-defined risk groups showed heterogeneity within each density category. Among fatty breasts, 76 of 237 examinations (32.1%) were classified as high risk and 90 (38.0%) as average risk. Among extremely dense breasts, 905 of 2,033 examinations (44.5%) were high risk and 540 (26.6%) were average risk. Compared with breast density alone, AI-defined risk categories further separated observed 5-year cancer incidence: incidence ranged from 2.1% to 3.6% across density categories alone, but from 0.0% to 5.8% after stratification by AI-defined risk category within each density group.

**Table 3.** Five-year cancer incidence stratified by breast imaging reporting and data system (BI-RADS) breast density category and AI-derived risk tier. The overall 5-year cancer incidence is provided for each BI-RADS density category (A–D). Within each density group, examinations are further subdivided into three AI risk tiers (average, intermediate, high) based on predicted 5-year risk, with the observed 5-year cancer incidence reported for each subgroup. Results are based on the subset of test-set examinations with sufficient follow-up for 5-year evaluation and available breast density information (n = 19,543).

| Breast Density N (%) | 5-Year Incidence | AI Risk Category | Risk Subgroup N (%) | Subgroup 5-Year Incidence |
| --- | --- | --- | --- | --- |
| A – Fatty 237 (1.2%) | 2.1% | High ($\geq 3.0\%$) | 76 (32.1%) | 5.3% |
| | | Intermediate ($\geq 1.7\%$ to $< 3.0\%$) | 71 (30.0%) | 1.4% |
| | | Average ($< 1.7\%$) | 90 (38.0%) | 0.0% |
| B – Scattered 4,749 (24.3%) | 2.7% | High ($\geq 3.0\%$) | 1,685 (35.5%) | 4.3% |
| | | Intermediate ($\geq 1.7\%$ to $< 3.0\%$) | 1,506 (31.7%) | 2.1% |
| | | Average ($< 1.7\%$) | 1,558 (32.8%) | 1.3% |
| C – Heterogeneous 12,524 (64.1%) | 3.6% | High ($\geq 3.0\%$) | 5,343 (42.7%) | 5.8% |
| | | Intermediate ($\geq 1.7\%$ to $< 3.0\%$) | 3,805 (30.4%) | 2.3% |
| | | Average ($< 1.7\%$) | 3,376 (27.0%) | 1.5% |
| D – Extremely Dense 2,033 (10.4%) | 2.9% | High ($\geq 3.0\%$) | 905 (44.5%) | 4.3% |
| | | Intermediate ($\geq 1.7\%$ to $< 3.0\%$) | 588 (28.9%) | 2.6% |
| | | Average ($< 1.7\%$) | 540 (26.6%) | 0.9% |

### Subgroup Analysis

Five-year AUCs across subgroups defined by age, breast density, and race are reported in Supplementary Table S4. Across racial subgroups, AUCs were 0.671 (95% CI, 0.576–0.756) for Black or African American patients, 0.650 (95% CI, 0.500–0.822) for Asian patients, 0.701 (95% CI, 0.400–0.864) for Hispanic patients, and 0.676 (95% CI, 0.645–0.706) for White patients. Across density groups, AUCs were 0.676 (95% CI, 0.614–0.735) for non-dense breasts and 0.678 (95% CI, 0.648–0.706) for dense breasts. Across age groups, AUCs ranged from 0.625 (95% CI, 0.566–0.683) among patients aged 70 years or older to 0.665 (95% CI, 0.616–0.715) among patients aged 50–59 years.

## Discussion

In this retrospective study of 122,072 women who underwent 295,298 breast ultrasound examinations, we developed BUS-Risk-Net to predict future breast cancer risk from breast ultrasound. Our results show that screening ultrasound carries risk-relevant signals. On the held-out test set, BUS-Risk-Net achieved higher discrimination performance than the reduced Tyrer–Cuzick model using age and ultrasound-estimated breast density (5-year AUC, 0.679 vs 0.594; *p* < 0.001). In an exploratory matched case-control analysis with complete risk-factor abstraction, BUS-Risk-Net also showed higher 5-year discrimination than the full Tyrer–Cuzick model (5-year AUC, 0.632 vs 0.514; *p* = 0.04). Moreover, BUS-Risk-Net also separated risk within each BI-RADS density category. Observed 5-year cancer incidence ranged from 0.0% to 5.3% in fatty breasts, 1.3% to 4.3% in scattered fibroglandular breasts, 1.5% to 5.8% in heterogeneously dense breasts, and 0.9% to 4.3% in extremely dense breasts.

Our results support the central hypothesis of this study that breast ultrasound images contain information relevant to future breast cancer risk. BUS-Risk-Net outperformed the TC model in the matched case-control cohort and the reduced Tyrer–Cuzick variant in the full held-out test set. The image-only model also retained predictive signal, indicating that ultrasound images contributed risk information beyond age and ultrasound-estimated breast density. This finding is consistent with prior studies showing that sonographic tissue features, including glandular tissue component, and background echotexture, are associated with breast cancer risk or established risk-related factors^22,23^.

Breast density is an established imaging marker of breast cancer risk. However, density alone provided limited separation of observed risk in our cohort: 5-year cancer incidence ranged from 2.1% to 3.6% across BI-RADS density categories. In contrast, after stratification by BUS-Risk-Net within each density category, observed incidence ranged more widely, from 0.0% to 5.8%. This pattern was present in both directions. Among women with extremely dense breasts, 540 of 2,033 examinations (26.6%) were classified as average risk, with an observed 5-year incidence of 0.9%. Conversely, among women with fatty breasts, 76 of 237 examinations (32.1%) were classified as high risk, with an observed 5-year incidence of 5.3%. These findings are consistent with current screening guidance that breast density should be considered together with overall risk, and they suggest that ultrasound-based AI may provide risk information beyond density alone.

Ultrasound-based risk prediction may serve different clinical roles across screening settings. In health systems where mammography is the primary screening modality, breast ultrasound is often used as supplemental imaging, particularly for women with dense breasts. Current ACR Appropriateness Criteria^40^ recommends that supplemental screening decisions consider both breast density and overall breast cancer risk, rather than breast density alone. Our findings support this approach. BUS-Risk-Net further stratified 5-year cancer incidence within each BI-RADS density category and identified lower-risk subgroups among women with dense breasts and higher-risk subgroups among women with non-dense breasts. As the model uses images from the supplemental ultrasound examination itself, it could provide an imaging-derived risk estimate at the time of care. After external and prospective validation, this information may help guide discussions about the need for future supplemental imaging and identify patients who may benefit from broader risk-reduction counseling, including formal risk assessment, genetic evaluation when appropriate, chemoprevention, or intensified screening. In limited-resource settings, where organized mammography may be difficult to implement, ultrasound is often used as an alternative imaging approach. In this setting, a risk estimate derived from the same screening ultrasound examination may be useful when mammography or detailed clinical risk-factor collection is not feasible. Prospective and external validation will be needed before clinical implementation.

Our work differs from most prior AI studies in breast ultrasound, which have focused primarily on cancer detection or lesion characterization rather than future cancer risk prediction. Existing studies have largely addressed lesion segmentation^41,42^, benign-versus-malignant classification^43^, and AI-assisted diagnostic interpretation to improve cancer detection and reduce false-positive findings or unnecessary biopsies^30^. Although some studies describe “risk stratification,” these approaches are typically diagnostic. They estimate the likelihood of malignancy in an existing lesion rather than the future risk of breast cancer in women without cancer at the index examination^44,45^. The most closely related work has used ultrasound to estimate breast density for incorporation into conventional clinical risk models^24^. In contrast, BUS-Risk-Net directly used the full screening ultrasound examination to predict 2- to 5-year breast cancer risk. This approach extends image-based risk prediction beyond mammography and digital breast tomosynthesis^10–12^ and suggests that screening ultrasound may contain risk-relevant imaging information beyond density alone. To our knowledge, BUS-Risk-Net is among the first models to directly predict long-term breast cancer risk from screening breast ultrasound examinations.

## Limitations

This study has several limitations. First, it was a single-institution retrospective study, and external validation is needed to assess generalizability across patient populations, practice settings, ultrasound systems, and acquisition protocols. Second, the cohort was predominantly White. Asian and Hispanic subgroups had smaller sample sizes and wider confidence intervals. Third, comparison with the full Tyrer–Cuzick model was limited to a small matched case-control cohort. Complete Tyrer–Cuzick risk factors were not routinely collected in clinical practice at our institution, and full Tyrer–Cuzick scores required labor-intensive manual chart review. Therefore, full Tyrer–Cuzick comparison could not be performed in the entire test set. Finally, BUS-Risk-Net was not evaluated prospectively, and this study did not assess whether model use would change clinical decisions, improve patient outcomes, reduce costs, or be acceptable to patients and clinicians. External and prospective validation will be needed before clinical implementation.

## Take Home Points

● BUS-Risk-Net enabled long-term breast cancer risk prediction directly from screening breast ultrasound.
● BUS-Risk-Net outperformed the evaluated Tyrer–Cuzick–based comparators in both the matched case–control cohort and the held-out screening test set.
● Within each breast density category, BUS-Risk-Net identified distinct risk groups with differing observed 5-year breast cancer incidence.

## Funding

This work was supported in part by the National Science Foundation (grant no. 1922658), the National Institutes of Health (grant no. 1R01EB036530-01A1 and 1UF1MH141129-01), the Milstein Pilot Project Fund, the Shifrin-Myers Breast Cancer Discovery Fund, and the Manhasset Women’s Coalition Against Breast Cancer.

## Author Contributions

Yuxuan Chen: Conceptualization, methodology, software, investigation, visualization, writing—original draft, writing—review and editing.

Haoyan Yang: Methodology, investigation, visualization, writing—review and editing. Yanqi Xu: Methodology, writing—review and editing.

Ria Soni: Data curation, validation, writing—review and editing.

Laura Heacock: Investigation, supervision, validation, clinical interpretation, writing—review and editing.

Maciej Lis: Validation, clinical interpretation, writing—review and editing. Aleksandra Stanek: Validation, clinical interpretation, writing—review and editing. Tomasz Puto: Validation, clinical interpretation, writing—review and editing.

Alana Amarosa Lewin: Clinical interpretation, supervision, writing—review and editing. Linda Moy: Clinical interpretation, supervision, writing—review and editing.

Freya R. Schnabel: Clinical interpretation, funding acquisition, project administration, supervision, writing—review and editing.

Yiqiu Shen: Conceptualization, methodology, supervision, funding acquisition, project administration, writing—review and editing.

## Data Availability

The data supporting the findings of this study contain protected health information and are not publicly available due to institutional and patient privacy restrictions.

## Acknowledgement

The authors thank Tedum Sampson for support with the computing environment; Luoyao Chen and Harold Stern for assistance with data retrieval; and Benny Huang for assistance with image extraction. Data from Institute A are proprietary and cannot be made publicly available. Source code for estimating breast density from ultrasound images is publicly available at https://github.com/hawaii-ai/bus-density.

## Notes

### Competing Interest Statement

The authors have declared no competing interest.

### Author Declarations

Ethics committee/IRB of NYU Langone Health gave ethical approval for this work

## References

1. Kim J, Harper A, McCormack V, et al. Global patterns and trends in breast cancer incidence and mortality across 185 countries. Nat Med. 2025;31(4):1154–1162. doi:10.1038/s41591-025-03502-3

2. Mendes J, Oliveira B, Araújo C, et al. Deep learning in breast cancer risk prediction: a review of recent applications in full-field digital mammography. Front Oncol. 2025;15(1656842):1656842. doi:10.3389/fonc.2025.1656842

3. Monticciolo DL, Newell MS, Moy L, Lee CS, Destounis SV. Breast cancer screening for women at higher-than-average risk: Updated recommendations from the ACR. J Am Coll Radiol. 2023;20(9):902–914. doi:10.1016/j.jacr.2023.04.002

4. Clift AK, Dodwell D, Lord S, et al. The current status of risk-stratified breast screening. Br J Cancer. 2022;126(4):533–550. doi:10.1038/s41416-021-01550-3

5. Gail MH, Brinton LA, Byar DP, et al. Projecting individualized probabilities of developing breast cancer for white females who are being examined annually. J Natl Cancer Inst. 1989;81(24):1879–1886. doi:10.1093/jnci/81.24.1879

6. Tice JA, Cummings SR, Smith-Bindman R, Ichikawa L, Barlow WE, Kerlikowske K. Using clinical factors and mammographic breast density to estimate breast cancer risk: development and validation of a new predictive model. Ann Intern Med. 2008;148(5):337–347. doi:10.7326/0003-4819-148-5-200803040-00004

7. Tice JA, Miglioretti DL, Li CS, Vachon CM, Gard CC, Kerlikowske K. Breast density and benign breast disease: Risk assessment to identify women at high risk of breast cancer. J Clin Oncol. 2015;33(28):3137–3143. doi:10.1200/JCO.2015.60.8869

8. Tyrer J, Duffy SW, Cuzick J. A breast cancer prediction model incorporating familial and personal risk factors. Stat Med. 2004;23(7):1111–1130. doi:10.1002/sim.1668

9. Arasu VA, Habel LA, Achacoso NS, et al. Comparison of mammography AI algorithms with a clinical risk model for 5-year breast cancer risk prediction: An observational study. Radiology. 2023;307(5):e222733. doi:10.1148/radiol.222733

10. Yala A, Mikhael PG, Strand F, et al. Toward robust mammography-based models for breast cancer risk. Sci Transl Med. 2021;13(578):eaba4373. doi:10.1126/scitranslmed.aba4373

11. Eriksson M, Destounis S, Czene K, et al. A risk model for digital breast tomosynthesis to predict breast cancer and guide clinical care. Sci Transl Med. 2022;14(644):eabn3971. doi:10.1126/scitranslmed.abn3971

12. Jiang S, Bennett DL, Colditz GA. Deriving a mammogram-based risk score from screening digital breast tomosynthesis for 5-year breast cancer risk prediction. Cancer Prev Res (Phila). 2025;18(6):347–354. doi:10.1158/1940-6207.CAPR-24-0427

13. Wang YK, Klanecek Z, Wagner T, et al. Using explainable AI to characterize features in the Mirai mammographic breast cancer risk prediction model. Radiol Artif Intell. 2025;7(6):e240417. doi:10.1148/ryai.240417

14. Donnelly J, Moffett L, Barnett AJ, et al. AsymMirai: Interpretable mammography-based deep learning model for 1-5-year breast cancer risk prediction. Radiology. 2024;310(3):e232780. doi:10.1148/radiol.232780

15. Dan Q, Zheng T, Liu L, Sun D, Chen Y. Ultrasound for Breast Cancer Screening in Resource-Limited Settings: Current Practice and Future Directions. Cancers. 2023;15(7):2112. doi:10.3390/cancers15072112

16. Sood R, Rositch AF, Shakoor D, et al. Ultrasound for breast cancer detection globally: A systematic review and meta-analysis. J Glob Oncol. 2019;5(5):1–17. doi:10.1200/JGO.19.00127

17. Kerlikowske K, Bissell MCS, Sprague BL, et al. Impact of BMI on prevalence of dense breasts by race and ethnicity. Cancer Epidemiol Biomarkers Prev. 2023;32(11):1524–1530. doi:10.1158/1055-9965.EPI-23-0049

18. Shen S, Zhou Y, Xu Y, et al. A multi-centre randomised trial comparing ultrasound vs mammography for screening breast cancer in high-risk Chinese women. Br J Cancer. 2015;112(6):998–1004. doi:10.1038/bjc.2015.33

19. Carney PA, Miglioretti DL, Yankaskas BC, et al. Individual and combined effects of age, breast density, and hormone replacement therapy use on the accuracy of screening mammography. Ann Intern Med. 2003;138(3):168–175. doi:10.7326/0003-4819-138-3-200302040-00008

20. Chotai N, Renganathan R, Uematsu T, et al. Breast cancer screening in Asian countries: Epidemiology, screening practices, outcomes, challenges, and future directions. Korean J Radiol. 2025;26(8):743–758. doi:10.3348/kjr.2025.0338

21. Wang FL, Chen F, Yin H, et al. Effects of age, breast density and volume on breast cancer diagnosis: a retrospective comparison of sensitivity of mammography and ultrasonography in China’s rural areas. Asian Pac J Cancer Prev. 2013;14(4):2277–2282. doi:10.7314/apjcp.2013.14.4.2277

22. Lee SH, Moon WK. Glandular tissue component on breast ultrasound in dense breasts: A new imaging biomarker for breast cancer risk. Korean J Radiol. 2022;23(6):574–580. doi:10.3348/kjr.2022.0099

23. Kim WH, Lee SH, Chang JM, Cho N, Moon WK. Background echotexture classification in breast ultrasound: inter-observer agreement study. Acta Radiol. 2017;58(12):1427–1433. doi:10.1177/0284185117695665

24. Bunnell A, Valdez D, Wolfgruber TK, et al. Prediction of mammographic breast density based on clinical breast ultrasound images using deep learning: a retrospective analysis. Lancet Reg Health Am. 2025;46(101096):101096. doi:10.1016/j.lana.2025.101096

25. Checklist for Artificial Intelligence in Medical Imaging (CLAIM). Accessed May 20, 2026. https://pubs.rsna.org/page/ai/claim

26. Park J, Witowski J, Xu Y, et al. A multi-modal AI system for screening mammography: Integrating 2D and 3D imaging to improve breast cancer detection in a prospective clinical study. *arXiv [eessIV]*. Published online April 7, 2025. doi:10.48550/arXiv.2504.05636

27. Tsue T, Mombourquette B, Taha A, Matthews TP, Vu YNT, Su J. Problems and shortcuts in deep learning for screening mammography. arXiv [csCV]. Published online March 28, 2023. doi:10.48550/arXiv.2303.16417

28. Lotter W, Diab AR, Haslam B, et al. Robust breast cancer detection in mammography and digital breast tomosynthesis using an annotation-efficient deep learning approach. Nat Med. 2021;27(2):244–249. doi:10.1038/s41591-020-01174-9

29. Shamout FE, Shen Y, Witowski J, et al. The NYU Breast Ultrasound Dataset v1.0. Published online 2021. https://cs.nyu.edu/~kgeras/reports/ultrasound_datav1.0.pdf

30. Shen Y, Shamout FE, Oliver JR, et al. Artificial intelligence system reduces false-positive findings in the interpretation of breast ultrasound exams. Nat Commun. 2021;12(1):5645. doi:10.1038/s41467-021-26023-2

31. Newell MS, Destounis SV, Leung JWT, DeMartini WB, Lee CH, Eby PR. ACR BI-RADS® v2025 Manual. *American College of Radiology, Reston*, VA. Published online 2025.

32. Jiao J, Zhou J, Li X, et al. USFM: A universal ultrasound foundation model generalized to tasks and organs towards label efficient image analysis. Med Image Anal. 2024;96(103202):103202. doi:10.1016/j.media.2024.103202

33. Ilse M, Tomczak J, Welling M. Attention-based Deep Multiple Instance Learning. In: Dy J, Krause A, eds. *Proceedings of the 35th International Conference on Machine Learning*. Vol 80. Proceedings of Machine Learning Research. PMLR; 10--15 Jul 2018:2127-2136. https://proceedings.mlr.press/v80/ilse18a.html

34. Platt J. Probabilistic outputs for support vector machines and comparisons to regularized likelihood methods. *Advances in large margin classifiers*. Published online 1999. https://www.researchgate.net/profile/John-Platt-2/publication/2594015_Probabilistic_Outputs_for_Support_Vector_Machines_and_Comparisons_to_Regularized_Likelihood_Methods/links/004635154cff5262d6000000/Probabilistic-Outputs-for-Support-Vector-Machines-and-Comparisons-to-Regularized-Likelihood-Methods.pdf

35. Cuzick J. IBIS Breast Cancer Risk Evaluation Tool. Accessed April 6, 2026. https://ems-trials.org/riskevaluator/

36. Jiang S, Bennett DL, Colditz GA. Validation of a dynamic risk prediction model incorporating prior mammograms in a diverse population. JAMA Netw Open. 2025;8(6):e2512681. doi:10.1001/jamanetworkopen.2025.12681

37. Lamb LR, Mercaldo SF, Ghaderi K, Carney A, Lehman CD. Comparison of the diagnostic accuracy of mammogram-based deep learning and traditional breast cancer risk models in patients who underwent supplemental screening with MRI. Radiology. 2023;308(3):e223077. doi:10.1148/radiol.223077

38. Bevers TB, Ward JH, Arun BK, et al. Breast cancer risk reduction, version 2.2015. J Natl Compr Canc Netw. 2015;13(7):880–915. doi:10.6004/jnccn.2015.0105

39. Visvanathan K, Fabian CJ, Bantug E, et al. Use of endocrine therapy for breast cancer risk reduction: ASCO clinical practice guideline update. J Clin Oncol. 2019;37(33):3152–3165. doi:10.1200/JCO.19.01472

40. Expert Panel on Breast Imaging, Paulis LV, Lewin AA, et al. ACR appropriateness criteria® supplemental breast cancer screening based on breast density: 2024 update. J Am Coll Radiol. 2025;22(5S):S405–S423. doi:10.1016/j.jacr.2025.02.023

41. Zhuang Z, Li N, Joseph Raj AN, Mahesh VGV, Qiu S. An RDAU-NET model for lesion segmentation in breast ultrasound images. PLoS One. 2019;14(8):e0221535. doi:10.1371/journal.pone.0221535

42. Pramanik P, Roy A, Cuevas E, Perez-Cisneros M, Sarkar R. DAU-Net: Dual attention-aided U-Net for segmenting tumor in breast ultrasound images. PLoS One. 2024;19(5):e0303670. doi:10.1371/journal.pone.0303670

43. Wang Y, Yao Y. Breast lesion detection using an anchor-free network from ultrasound images with segmentation-based enhancement. Sci Rep. 2022;12(1):14720. doi:10.1038/s41598-022-18747-y

44. Gu Y, Xu W, Liu T, et al. Ultrasound-based deep learning in the establishment of a breast lesion risk stratification system: a multicenter study. Eur Radiol. 2023;33(4):2954–2964. doi:10.1007/s00330-022-09263-8

45. Liu T, An X, Liu Y, et al. A novel deep learning system for breast lesion risk stratification in ultrasound images. In: Lecture Notes in Computer Science. Lecture Notes in Computer Science. Springer Nature Switzerland; 2022:472–481. doi:10.1007/978-3-031-16437-8_45

