## Supplementary Materials for "Five-Year Breast Cancer Risk Prediction From Screening Breast Ultrasound Using Deep Learning"

**Supplementary Table 1.** Characteristics of the matched case–control cohort. Cases and controls were matched 1:1 by age group, race, and breast density. Breast density was categorized according to the breast imaging reporting and data system (BI-RADS) as non-dense (BI-RADS A–B) or dense (BI-RADS C–D).

| Characteristic | Category | Cases | Controls |
| --- | --- | --- | --- |
| Total | All | 120 | 120 |
| Age group | <40 | 3 (2.5%) | 3 (2.5%) |
| | 40–49 | 22 (18.3%) | 22 (18.3%) |
| | 50–59 | 35 (29.2%) | 35 (29.2%) |
| | 60–69 | 37 (30.8%) | 37 (30.8%) |
| | ≥70 | 23 (19.2%) | 23 (19.2%) |
| Density | Non-dense (BI-RADS A–B) | 30 (25.0%) | 30 (25.0%) |
| | Dense (BI-RADS C–D) | 90 (75.0%) | 90 (75.0%) |
| Race | White | 102 (85.0%) | 102 (85.0%) |
| | African American | 12 (10.0%) | 12 (10.0%) |
| | Asian | 4 (3.3%) | 4 (3.3%) |
| | Hispanic | 2 (1.7%) | 2 (1.7%) |

**Supplementary Table 2.** Distribution of time to breast cancer diagnosis across data splits. Values represent the number of examinations followed by a biopsy-confirmed breast cancer diagnosis within each time interval after the index ultrasound examination. Percentages are calculated relative to the total number of cancer cases within each split.

| Time to Cancer | Training Set, n (%) | Validation Set, n (%) | Test Set, n (%) |
|---|---|---|---|
| 0.5–1 year | 39 (0.9%) | 6 (1.4%) | 3 (0.5%) |
| 1–2 years | 672 (14.9%) | 61 (13.9%) | 80 (12.5%) |
| 2–3 years | 1,301 (28.9%) | 128 (29.2%) | 206 (32.1%) |
| 3–4 years | 1,278 (28.4%) | 120 (27.4%) | 175 (27.3%) |
| 4–5 years | 1,210 (26.9%) | 123 (28.1%) | 178 (27.7%) |
| Total cancer cases | 4,500 (100%) | 438 (100%) | 642 (100%) |

**Supplementary Table 3.** Number of examinations and unique patients evaluated at each prediction horizon in the held-out test set. Positive examinations were followed by a biopsy-confirmed breast cancer diagnosis within the specified horizon; negative examinations had sufficient follow-up confirming the absence of malignancy beyond that horizon. Counts are reported for 2-, 3-, 4-, and 5-year risk prediction.

| Prediction Horizon | Positive Examinations | Negative Examinations | Total Evaluated Examinations | Unique Patients |
|---|---|---|---|---|
| 2-Year | 83 | 30,557 | 30,640 | 12,010 |
| 3-Year | 289 | 28,318 | 28,607 | 11,291 |
| 4-Year | 464 | 24,117 | 24,581 | 10,297 |
| 5-Year | 642 | 18,906 | 19,548 | 9,015 |

**Supplementary Table 4.** Five-year breast cancer risk prediction performance across subgroups in the held-out test set. Examination-level analyses are reported as areas under the receiver operating characteristic curve (AUCs) with 95% confidence intervals (CIs). Age analyses included all 5-year evaluable examinations (n = 19,548); race and breast density analyses included examinations with available race (n = 16,056) and density (n = 19,543) information, respectively. Breast density was categorized as non-dense (BI-RADS A–B) or dense (BI-RADS C–D)

| Subgroup Type | Subgroup | n | Positive (n) | AUC |
|---|---|---|---|---|
| Race | African American | 1,488 | 56 | 0.671 (0.576–0.756) |
| | Asian | 594 | 19 | 0.650 (0.500–0.822) |
| | Hispanic | 179 | 3 | 0.701 (0.400–0.864) |
| | White | 13,795 | 503 | 0.676 (0.645–0.706) |
| Age | <40 | 363 | 9 | 0.632 (0.477–0.766) |
| | 40–49 | 4,472 | 99 | 0.654 (0.585–0.720) |
| | 50–59 | 6,602 | 165 | 0.665 (0.616–0.715) |
| | 60–69 | 5,569 | 208 | 0.654 (0.602–0.705) |
| | ≥70 | 2,542 | 161 | 0.625 (0.566–0.683) |
| Density | Non-dense (BI-RADS A-B) | 4,986 | 131 | 0.676 (0.614–0.735) |
| | Dense (BI-RADS C-D) | 14,557 | 511 | 0.678 (0.648–0.706) |

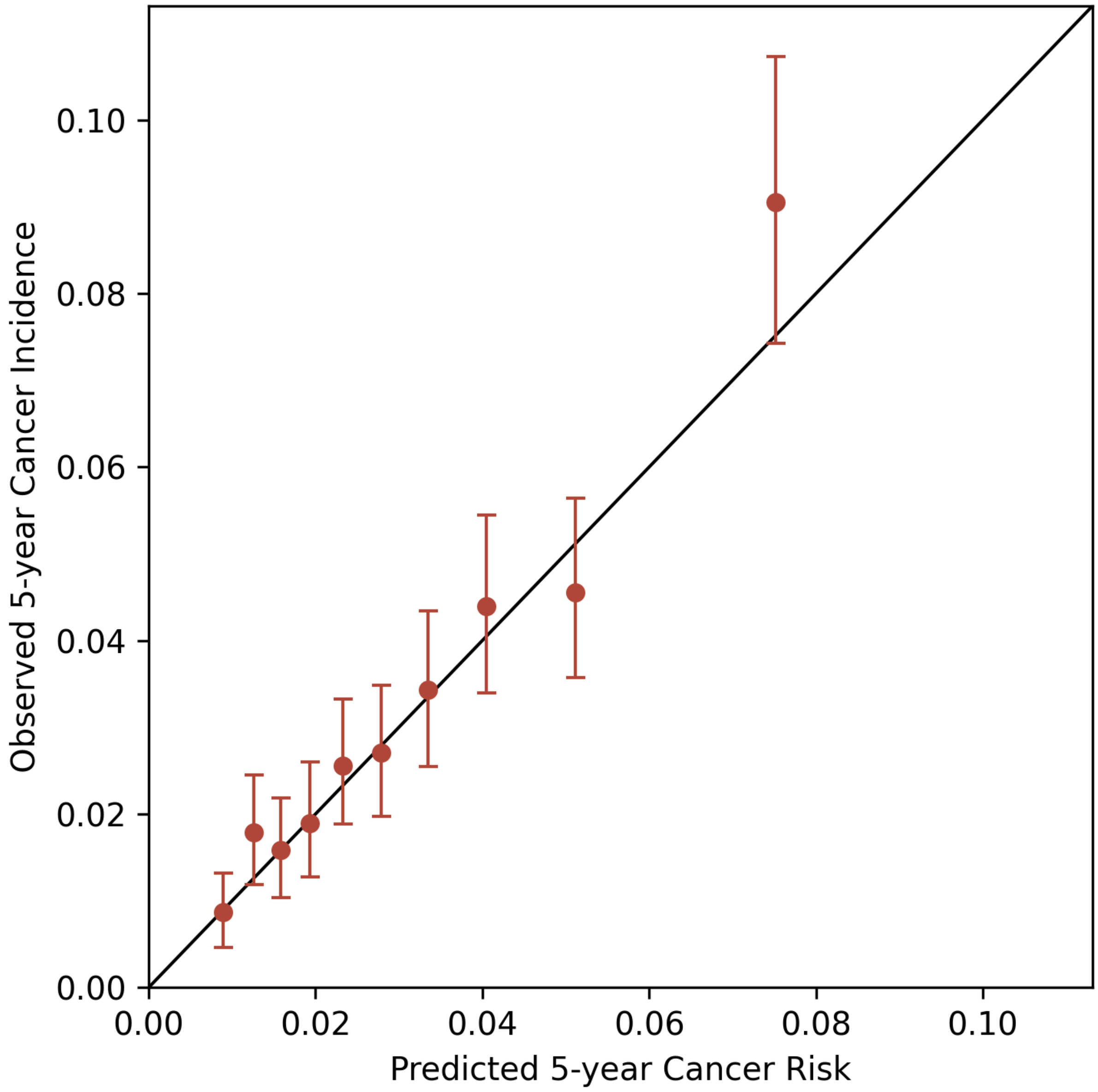


**Supplementary Figure 1.** Calibration plot for 5-year breast cancer risk in the held-out test set. The plot shows observed 5-year breast cancer incidence versus predicted risk across deciles of predicted risk, with 95% confidence intervals (CIs) for the observed incidence. The dashed line indicates perfect calibration.
